# Inequalities in healthcare disruptions during the Covid-19 pandemic: Evidence from 12 UK population-based longitudinal studies

**DOI:** 10.1101/2021.06.08.21258546

**Authors:** Jane Maddock, Sam Parsons, Giorgio Di Gessa, Michael J. Green, Ellen J. Thompson, Anna J. Stevenson, Alex S.F. Kwong, Eoin McElroy, Gillian Santorelli, Richard J. Silverwood, Gabriella Captur, Nish Chaturvedi, Claire J. Steves, Andrew Steptoe, Praveetha Patalay, George B. Ploubidis, Srinivasa Vittal Katikireddi

## Abstract

**Background:** Health systems worldwide have faced major disruptions due to COVID-19 which could exacerbate health inequalities. The UK National Health Service (NHS) provides free healthcare and prioritises equity of delivery, but the pandemic may be hindering the achievement of these goals. We investigated associations between multiple social characteristics (sex, age, occupational social class, education and ethnicity) and self-reported healthcare disruptions in over 65,000 participants across twelve UK longitudinal studies.

**Methods:** Participants reported disruptions from March 2020 up to late January 2021. Associations between social characteristics and three types of self-reported healthcare disruption (medication access, procedures, appointments) and a composite of any of these were assessed in logistic regression models, adjusting for age, sex and ethnicity where relevant. Random-effects meta-analysis was conducted to obtain pooled estimates.

**Results:** Prevalence of disruption varied across studies; between 6.4% (TwinsUK) and 31.8 % (Understanding Society) of study participants reported any disruption. Females (Odd Ratio (OR): 1.27 [95%CI: 1.15,1.40]; I^2^=53%), older persons (e.g. OR: 1.39 [1.13,1.72]; I^2^=77% for 65-75y vs 45-54y), and Ethnic minorities (excluding White minorities) (OR: 1.19 [1.05,1.35]; I^2^=0% vs White) were more likely to report healthcare disruptions. Those in a more disadvantaged social class (e.g. OR: 1.17 [1.08, 1.27]; I^2^=0% for manual/routine vs managerial/professional) were also more likely to report healthcare disruptions, but no clear differences were observed by education levels.

**Conclusion:** The COVID-19 pandemic has led to unequal healthcare disruptions, which, if unaddressed, could contribute to the maintenance or widening of existing health inequalities.

## Introduction

The coronavirus disease 2019 (COVID-19) pandemic is profoundly affecting all aspects of society. Health systems world-wide have faced major disruption as they respond to large increases in demand arising from the COVID-19 disease.^1–4^ Furthermore, healthcare access has been reduced by governmental control measures and the public’s fear of contracting infection.^5^ Disruptions may have both short and long-term health consequences as preventive treatments are foregone, disease surveillance is interrupted and disease diagnoses are delayed. While the disruption of health systems can impact the entire population, it has become apparent that not all groups have been affected equally. Understanding the impacts of the pandemic on health systems and on equity of healthcare access is therefore a major policy priority.

In the UK, the National Health Service (NHS) provides free healthcare and prioritises equity of delivery. However, the UK’s relatively high COVID-19 burden and associated repeated lockdown measures have raised concerns that the health system may not be providing accessible care to those who need it most. Recent reports from NHS Digital indicate a large increase in those waiting 12 months or more for elective treatments in February 2021 compared to March 2020.^6^ Furthermore, despite decreases in attendance at accident and emergency services,^4^ the number of patients waiting over 12 hours for admission was 34% higher in January 2021 than January 2020. Disruption to pharmacological treatments has also been reported with delays in accessing medication.^7,8^ However, a comprehensive assessment of inequalities in healthcare disruption in the community is lacking.

It is well known that health systems do not meet the needs of all social groups equitably, with marked health inequalities by sex, ethnicity, and socioeconomic position.^9,10^ For example, the inverse care law demonstrates that health service provision is often not allocated according to need, with more socioeconomically deprived areas relatively under-served.^11^ Given the barriers that some social groups face in accessing high quality healthcare, there is considerable concern that disadvantaged groups (e.g., ethnic minorities) will be disproportionately impacted by healthcare disruption during the COVID-19 pandemic, as some emerging evidence suggests.^12,13^

Harnessing multiple longitudinal studies allows inequalities to be studied in detail by improving statistical power and allows consistency of findings to be investigated. We therefore aimed to investigate inequalities in healthcare disruption during the COVID-19 pandemic in 12 population-based longitudinal studies, to help inform targeting of policy responses as we move out of the acute phase of the pandemic. In particular, we investigate healthcare disruptions (including prescription or medication access, procedures or surgery, clinical appointments) by sex, age, ethnicity, education, and occupational social class and we explore whether associations differ by age, or for those who have been recommended to ‘shield’ due to clinical vulnerability.

## Methods

### Design

The UK National Core Studies – Longitudinal Health and Wellbeing programme aims to draw together data from multiple UK population-based longitudinal studies to answer questions relevant to the pandemic response. By coordinating analyses within each study and statistically pooling results in a meta-analysis, we can provide robust evidence to understand how the pandemic has impacted population health and support efforts to mitigate its health effects going forward.

### Participants

Data here were from 12 UK population studies which had conducted surveys both before and during the COVID-19 pandemic. Details of the design, sample frames, current age range, timing of the COVID-19 surveys, response rates, and analytical sample size are available in Supplementary Table S1. Ethics statement and data access details can be found in Supplementary Table S2.

Six of these studies were age homogenous birth cohorts (where all individuals were of similar age within each cohort): the Millennium Cohort Study (MCS);^14^ the Avon Longitudinal Study of Parents and Children (ALSPAC-G1);^15^ Next Steps (NS);^16^ the 1970 British Cohort Study (BCS);^17^ the National Child Development Study (NCDS);^18^ and the National Survey of Health and Development (NSHD).^19,20^ Six further age heterogeneous studies (each covering a range of age groups) were included: the Born in Bradford study (BIB);^21,22^ Understanding Society (USOC);^23^ Generation Scotland: the Scottish Family Health Study (GS);^24^ the parents of the ALSPAC-G1 cohort which we refer to as ALSPAC-G0;^25^ the UK Adult Twin Registry (TwinsUK);^26,27^ and the English Longitudinal Study of Ageing (ELSA).^28^ Analytical samples were defined within each study based on those who had recorded at least one healthcare disruption outcome in a COVID-19 survey and had valid data on a minimum set of covariates (sex, ethnicity, and age where relevant). Most studies were weighted to be representative of their target populations accounting for differential non-response.^20,29,30^ Weights were not available for BIB or TwinsUK. Studies were ordered for presentation by age of sample (youngest to oldest), with the age homogenous cohorts first, followed by the age heterogenous studies.

### Measures

Below we describe the overall approach to measuring each variable in the analysis. Full details of the questions and coding used within each cohort are available in Supplementary File 1.

### Outcomes

We assessed self-reported disruptions to: prescriptions or medication access; procedures or surgery; and appointments (e.g., with a GP or outpatient services); and a combined variable indicating disruptions to any of the afore-mentioned. Any deviation from planned/existing treatment was coded as a disruption, regardless of the reason for the disruption. Where multiple pandemic survey waves had been included, we coded for any disruptions reported up to and including the most recent. This meant at least 7 months of follow-up for most studies (GS had five and ELSA four, while ALSPAC had the longest follow-up period at nine months).

### Indicators of Inequality

We assessed inequalities associated with key sociodemographic characteristics, i.e. sex, age, ethnicity, education, and occupational social class. For age, we considered age-groups categorised as: 16-24; 25-34; 35-44; 45-54; 55-64; 65-74; and 75+ years. Depending on the ethnic variation of each study and on the level of detail available, we examined both a binary (White [including White minorities] vs Ethnic minorities [excluding White minorities]) and a finer categorisation of ethnicity (White, South Asian, Black, Mixed, Other Asian, Other Ethnic Minority). For education we distinguished between degree or equivalent; A-level or equivalent (i.e. post-compulsory schooling qualifications); GCSE or equivalent (i.e. qualifications for completing compulsory schooling); and fewer or no qualifications. We also examined occupational class with the following categories (based on different coding schemes in different studies): Professional/Managerial; Intermediate; Routine/Manual; and Other (which included never/long-term non-employed and, in some studies, respondents who could not be classified elsewhere). Where respondents’ education and occupational class were not available, we considered parental education or household social class. For full details, see Supplementary File 1.

### Moderators

We decided *a priori* to examine modification by age and clinical vulnerability to COVID-19 to see whether inequalities varied by life-stage or were particularly acute for those with higher healthcare needs and at higher risk from COVID-19 harms. For moderation by age, the age-heterogeneous studies split their samples into the age-bands covered, while age homogeneous cohorts were included within the appropriate age bands (see above for banding). In the UK, clinically extremely vulnerable people were advised to stay at home (‘shield’) during the pandemic. In this paper, shielding status was based on whether respondents reported having received a letter from the NHS advising them to stay at home to protect themselves.

### Other variables

The following covariates were also included where relevant and available within each study: UK Nation (i.e. England, Scotland, Wales, or Northern Ireland); household composition (based on partnership status and whether there were children in the household); and pre-pandemic self-reported health (good vs poor).

## Analysis

Within each study, distributions of sociodemographic characteristics and healthcare disruption were examined. Then, each healthcare disruption outcome was regressed on each indicator of inequality (i.e. sex, age, ethnicity, education, and occupational class). Unadjusted associations are included in Supplementary File 2. Since our aim was primarily to describe inequalities, we focus on presenting associations with minimal adjustment only for sex, age, and ethnicity when applicable. To assess whether associations were independent of other related factors, we also provide supplementary results for any healthcare disruption which additionally adjust for: education, occupational class, UK Nation (where appropriate), household composition, and pre-pandemic self-reported health. Moderation by age and shielding status was assessed using stratified models.

Results were then meta-analysed for each outcome for the full sample, and within age and shielding strata. We used a random effects meta-analysis with restricted maximum likelihood. We report heterogeneity using the I^2^ statistic (0% indicates low variation between estimates across studies, while values closer to 100% indicate greater heterogeneity). Meta-analyses were conducted in Stata 16 (V.16).^31^

## Results

### Descriptive Statistics

The distribution of demographic and socio-economic characteristics within each study is presented in Table 1. A total of 68,912 participants were included in the coordinated analysis. Participants from BIB were all female, as were the vast majority (89.4%) from TwinsUK. The mean age ranged from 16 years in BIB and USOC to 90+ years in TwinsUK and ELSA.

**Table 1.**
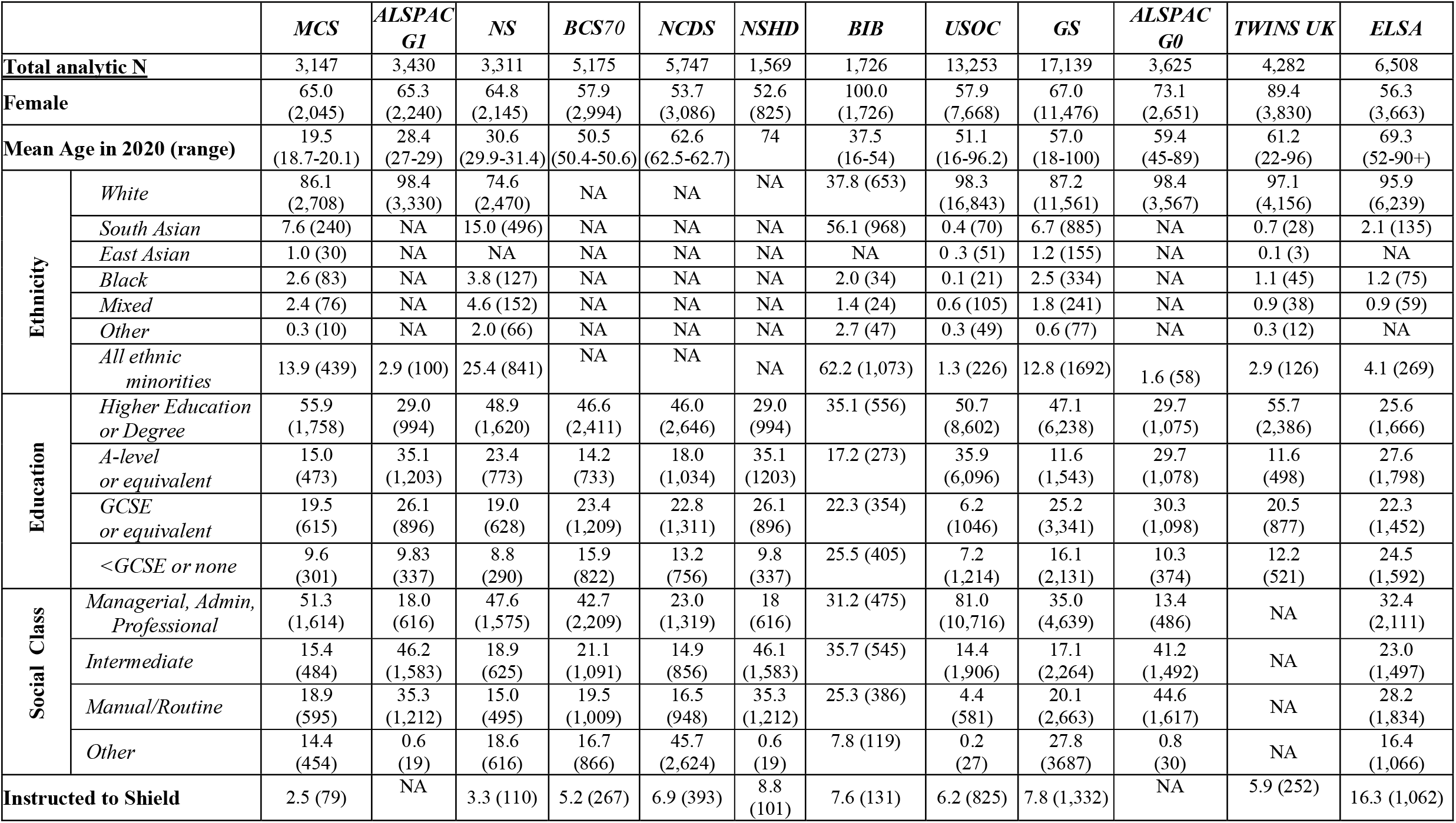
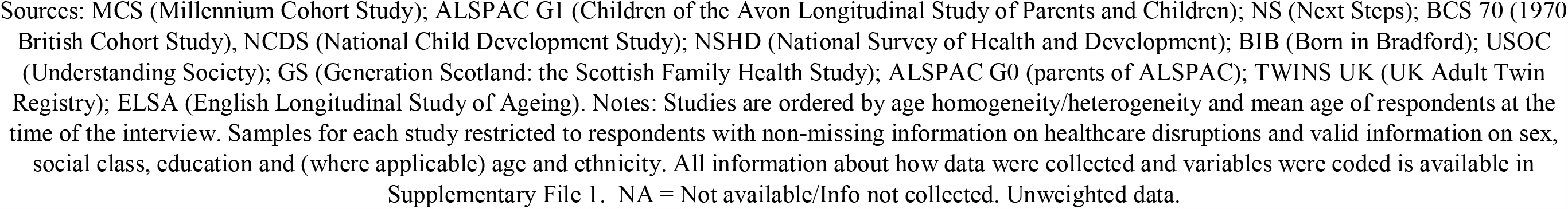
Percent (and N) distribution of demographic and socio-economic characteristics by study.

|  |  | *MCS* | *ALSPAC*<br>*G1* | *NS* | *BCS70* | *NCDS* | *NSHD* | *BIB* | *USOC* | *GS* | *ALSPAC*<br>*G0* | *TWINS UK* | *ELSA* |
| --- | --- | --- | --- | --- | --- | --- | --- | --- | --- | --- | --- | --- | --- |
| **Total analytic N** |  | 3,147 | 3,430 | 3,311 | 5,175 | 5,747 | 1,569 | 1,726 | 13,253 | 17,139 | 3,625 | 4,282 | 6,508 |
| **Female** |  | 65.0<br>(2,045) | 65.3<br>(2,240) | 64.8<br>(2,145) | 57.9<br>(2,994) | 53.7<br>(3,086) | 52.6<br>(825) | 100.0<br>(1,726) | 57.9<br>(7,668) | 67.0<br>(11,476) | 73.1<br>(2,651) | 89.4<br>(3,830) | 56.3<br>(3,663) |
| **Mean Age in 2020 (range)** |  | 19.5<br>(18.7-20.1) | 28.4<br>(27-29) | 30.6<br>(29.9-31.4) | 50.5<br>(50.4-50.6) | 62.6<br>(62.5-62.7) | 74 | 37.5<br>(16-54) | 51.1<br>(16-96.2) | 57.0<br>(18-100) | 59.4<br>(45-89) | 61.2<br>(22-96) | 69.3<br>(52-90+) |
| **Ethnicity** | *White* | 86.1<br>(2,708) | 98.4<br>(3,330) | 74.6<br>(2,470) | NA | NA | NA | 37.8 (653) | 98.3<br>(16,843) | 87.2<br>(11,561) | 98.4<br>(3,567) | 97.1<br>(4,156) | 95.9<br>(6,239) |
|  | *South Asian* | 7.6 (240) | NA | 15.0 (496) | NA | NA | NA | 56.1 (968) | 0.4 (70) | 6.7 (885) | NA | 0.7 (28) | 2.1 (135) |
|  | *East Asian* | 1.0 (30) | NA | NA | NA | NA | NA | NA | 0.3 (51) | 1.2 (155) | NA | 0.1 (3) | NA |
|  | *Black* | 2.6 (83) | NA | 3.8 (127) | NA | NA | NA | 2.0 (34) | 0.1 (21) | 2.5 (334) | NA | 1.1 (45) | 1.2 (75) |
|  | *Mixed* | 2.4 (76) | NA | 4.6 (152) | NA | NA | NA | 1.4 (24) | 0.6 (105) | 1.8 (241) | NA | 0.9 (38) | 0.9 (59) |
|  | *Other* | 0.3 (10) | NA | 2.0 (66) | NA | NA | NA | 2.7 (47) | 0.3 (49) | 0.6 (77) | NA | 0.3 (12) | NA |
|  | *All ethnic minorities* | 13.9 (439) | 2.9 (100) | 25.4 (841) | NA | NA | NA | 62.2 (1,073) | 1.3 (226) | 12.8 (1692) | 1.6 (58) | 2.9 (126) | 4.1 (269) |
| **Education** | *Higher Education or Degree* | 55.9<br>(1,758) | 29.0<br>(994) | 48.9<br>(1,620) | 46.6<br>(2,411) | 46.0<br>(2,646) | 29.0<br>(994) | 35.1 (556) | 50.7<br>(8,602) | 47.1<br>(6,238) | 29.7<br>(1,075) | 55.7<br>(2,386) | 25.6<br>(1,666) |
|  | *A-level or equivalent* | 15.0<br>(473) | 35.1<br>(1,203) | 23.4<br>(773) | 14.2<br>(733) | 18.0<br>(1,034) | 35.1<br>(1203) | 17.2 (273) | 35.9<br>(6,096) | 11.6<br>(1,543) | 29.7<br>(1,078) | 11.6<br>(498) | 27.6<br>(1,798) |
|  | *GCSE or equivalent* | 19.5<br>(615) | 26.1<br>(896) | 19.0<br>(628) | 23.4<br>(1,209) | 22.8<br>(1,311) | 26.1<br>(896) | 22.3 (354) | 6.2<br>(1046) | 25.2<br>(3,341) | 30.3<br>(1,098) | 20.5<br>(877) | 22.3<br>(1,452) |
|  | *<GCSE or none* | 9.6<br>(301) | 9.83<br>(337) | 8.8<br>(290) | 15.9<br>(822) | 13.2<br>(756) | 9.8<br>(337) | 25.5 (405) | 7.2<br>(1,214) | 16.1<br>(2,131) | 10.3<br>(374) | 12.2<br>(521) | 24.5<br>(1,592) |
| **Social Class** | *Managerial, Admin, Professional* | 51.3<br>(1,614) | 18.0<br>(616) | 47.6<br>(1,575) | 42.7<br>(2,209) | 23.0<br>(1,319) | 18<br>(616) | 31.2 (475) | 81.0<br>(10,716) | 35.0<br>(4,639) | 13.4<br>(486) | NA | 32.4<br>(2,111) |
|  | *Intermediate* | 15.4<br>(484) | 46.2<br>(1,583) | 18.9<br>(625) | 21.1<br>(1,091) | 14.9<br>(856) | 46.1<br>(1,583) | 35.7 (545) | 14.4<br>(1,906) | 17.1<br>(2,264) | 41.2<br>(1,492) | NA | 23.0<br>(1,497) |
|  | *Manual/Routine* | 18.9<br>(595) | 35.3<br>(1,212) | 15.0<br>(495) | 19.5<br>(1,009) | 16.5<br>(948) | 35.3<br>(1,212) | 25.3 (386) | 4.4<br>(581) | 20.1<br>(2,663) | 44.6<br>(1,617) | NA | 28.2<br>(1,834) |
|  | *Other* | 14.4<br>(454) | 0.6<br>(19) | 18.6<br>(616) | 16.7<br>(866) | 45.7<br>(2,624) | 0.6<br>(19) | 7.8 (119) | 0.2<br>(27) | 27.8<br>(3687) | 0.8<br>(30) | NA | 16.4<br>(1,066) |
| **Instructed to Shield** |  | 2.5 (79) | NA | 3.3 (110) | 5.2 (267) | 6.9 (393) | 8.8<br>(101) | 7.6 (131) | 6.2 (825) | 7.8 (1,332) | NA | 5.9 (252) | 16.3 (1,062) |

Sources: MCS (Millennium Cohort Study); ALSPAC G1 (Children of the Avon Longitudinal Study of Parents and Children); NS (Next Steps); BCS 70 (1970 British Cohort Study), NCDS (National Child Development Study); NSHD (National Survey of Health and Development); BIB (Born in Bradford); USOC (Understanding Society); GS (Generation Scotland: the Scottish Family Health Study); ALSPAC G0 (parents of ALSPAC); TWINS UK (UK Adult Twin Registry); ELSA (English Longitudinal Study of Ageing). Notes: Studies are ordered by age homogeneity/heterogeneity and mean age of respondents at the time of the interview. Samples for each study restricted to respondents with non-missing information on healthcare disruptions and valid information on sex, social class, education and (where applicable) age and ethnicity. All information about how data were collected and variables were coded is available in Supplementary File 1. NA = Not available/Info not collected. Unweighted data.

Overall, the prevalence of any healthcare disruption ranged from 6.4% in TwinsUK to 31.8% in USOC (Figure 1). Table 2 shows that disruptions to medical appointments were most common, ranging from 3.5% (ELSA) to 28.4% (USOC). This was followed by disruptions in prescriptions or medication access, varying from 1.2% (BIB) to 10.4% (GS). Disruptions to procedures or surgery were least common ranging from 0.7% (MCS) to 16.8% (ELSA).

**Table 2.** Percent prevalence (and 95% confidence intervals) of healthcare disruptions during the pandemic, by study.

|  | *MCS* | *ALSPAC (G1)* | *NS* □ | *BCS70* | *NCDS* | *NSHD* | *BIB* | *USOC* | *GS* | *ALSPAC (G0)* □ | *TWINS UK* | *ELSA* |
| --- | --- | --- | --- | --- | --- | --- | --- | --- | --- | --- | --- | --- |
| **Prescription/ medication access** | 4.0<br>(2.3-5.5) | NA | 3.8<br>(2.3-5.3) | 3.4<br>(2.7-4.2) | 2.4<br>(1.8-3.0) | 2.2<br>(1.3-3.8) | 1.2<br>(0.7-1.7) | 5.5<br>(5.0-6.1) | 10.4<br>(9.9-10.9) | NA | 2.9<br>(2.5-3.3) | 0.8<br>(0.6-1.2) |
| **Procedures or surgery** | 0.7<br>(0.0-1.2) | 1.6<br>(1.2-2.1) | 2.1<br>(0.0-3.8) | 1.0<br>(0.7-1.2) | 2.8<br>(2.0-3.5) | 2.5<br>(1.4-4.4) | NA | 12.3<br>(11.6-13.0) | 2.1<br>(1.9-2.4) | 2.9<br>(2.1-3.9) | NA | 16.8<br>(15.7-17.9) |
| **Appointments** | 6.2<br>(4.9-7.6) | 11.7<br>(10.3-13.2) | 7.3<br>(5.6-9.0) | 10.6<br>(9.2-12.1) | 12.1<br>(10.9-13.3) | 12.0<br>(9.3-15.6) | 8.6<br>(7.4-10.1) | 28.4<br>(27.4-29.4) | 16.6<br>(16.0-17.1) | 14.4<br>(12.8-16.2) | NA | 3.5<br>(2.9-4.1) |
| **Any healthcare disruption** | 10.1<br>(8.1-12.1) | 15.9<br>(14.3-17.6) | 12.8<br>(10.3-15.4) | 14.3<br>(12.7-15.9) | 16.7<br>(15.2-18.2) | 16.4<br>(13.2-20.2) | 9.4<br>(8.1-10.9) | 31.8<br>(30.8-32.8) | 25.3<br>(24.6-25.9) | 19.9<br>(18.1-21.9) | 6.35<br>(5.9-7.2) | 19.5<br>(18.3-20.8) |

Sources: MCS (Millennium Cohort Study); ALSPAC G1 (Children of the Avon Longitudinal Study of Parents and Children); NS (Next Steps); BCS 70 (1970 British Cohort Study); NCDS (National Child Development Study); NSHD (National Survey of Health and Development); BIB (Born in Bradford); USOC (Understanding Society); GS (Generation Scotland: the Scottish Family Health Study); ALSPAC G0 (parents of ALSPAC); TWINS UK (UK Adult Twin Registry); ELSA (English Longitudinal Study of Ageing). Notes: Studies are ordered by age homogeneity/heterogeneity and mean age of respondents at the time of the interview. Samples for each study restricted to respondents with non-missing information on healthcare disruptions and valid information on sex, social class, education and (where applicable) age and ethnicity. All information about how data were collected and variables were coded is available in Supplementary File 1. TWINSUK had an additional question: “Have you experienced healthcare disruption as a result of the COVID-19 pandemic?” This data was also used to derive the ‘any healthcare disruption’ variable for TWINSUK. NA = Not available/Info not collected. Weighted data where applicable

**Figure 1.**
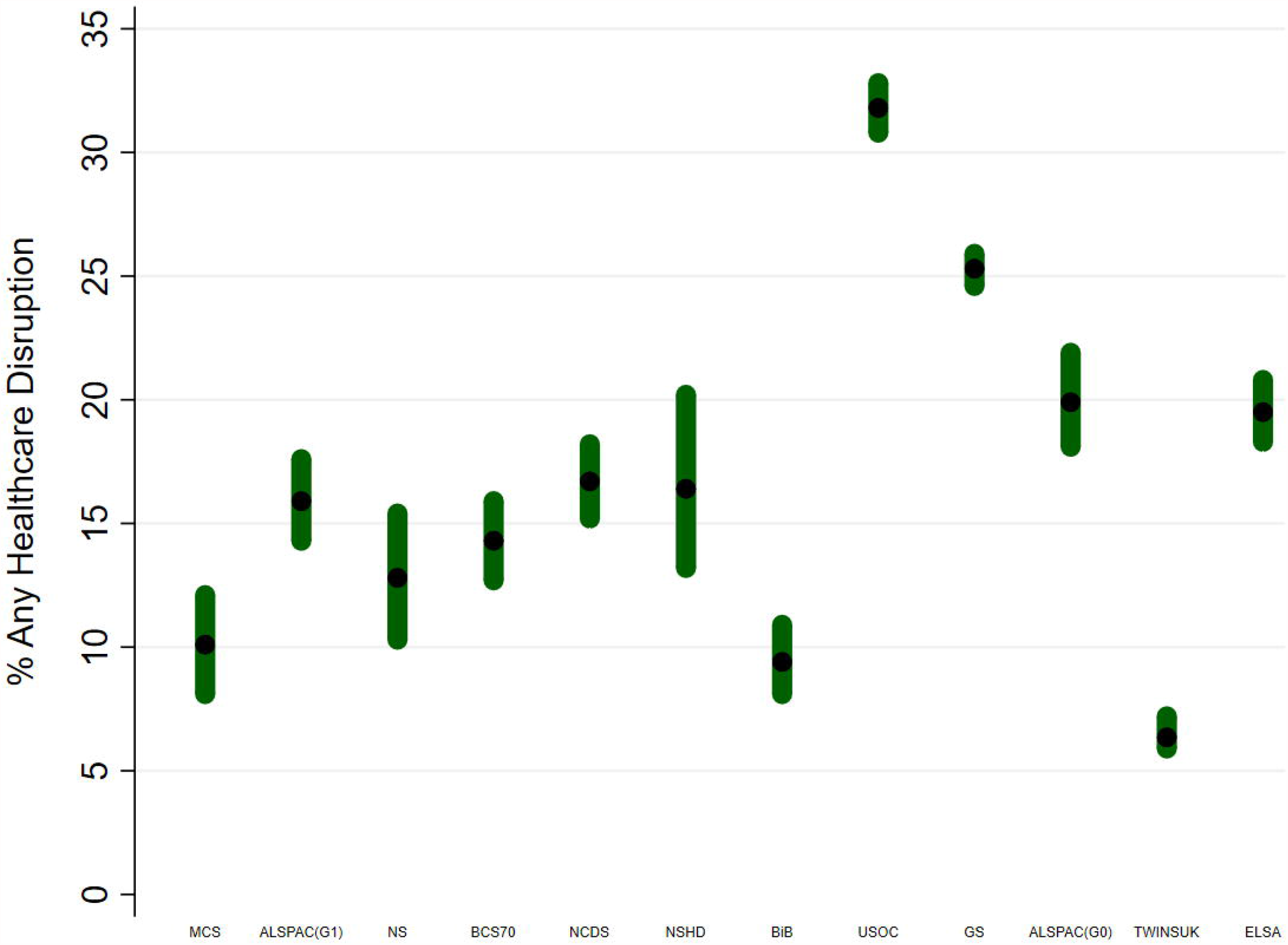
Prevalence (and 95% CIs) of any healthcare disruption by study. Sources: MCS (Millennium Cohort Study); ALSPAC G1 (Children of the Avon Longitudinal Study of Parents and Children); NS (Next Steps); BCS 70 (1970 British Cohort Study), NCDS (National Child Development Study); NSHD (National Survey of Health and Development); BIB (Born in Bradford); USOC (Understanding Society); GS (Generation Scotland: the Scottish Family Health Study); ALSPAC G0 (parents of ALSPAC); TWINS UK (UK Adult Twin Registry); ELSA (English Longitudinal Study of Ageing). Notes: Studies are ordered by age homogeneity/heterogeneity and mean age of respondents at the time of the interview. Samples for each study restricted to respondents with non-missing information on healthcare disruptions and valid information on sex, social class, education and (where applicable) age and ethnicity. All information about how data were collected and variables were coded is available in Supplementary File 1.

### Sex and healthcare disruptions

Across all studies females were generally more likely to report any healthcare disruptions than males (Supplementary Table S3 for details).

Pooled results from the meta-analysis demonstrate that females had increased odds of any healthcare disruption compared with males (OR: 1.27 [95%CI: 1.15,1.40]; I^2^=54%, figure 2, supplementary file 2). Similar associations were observed for disruptions to appointments (OR: 1.33 [95%CI: 1.17,1.52]; I^2^=60%). The association between sex and the less prevalent disruptions to procedures and medications crossed the null (Supplementary File 2 and Figure 2).

**Figure 2:**
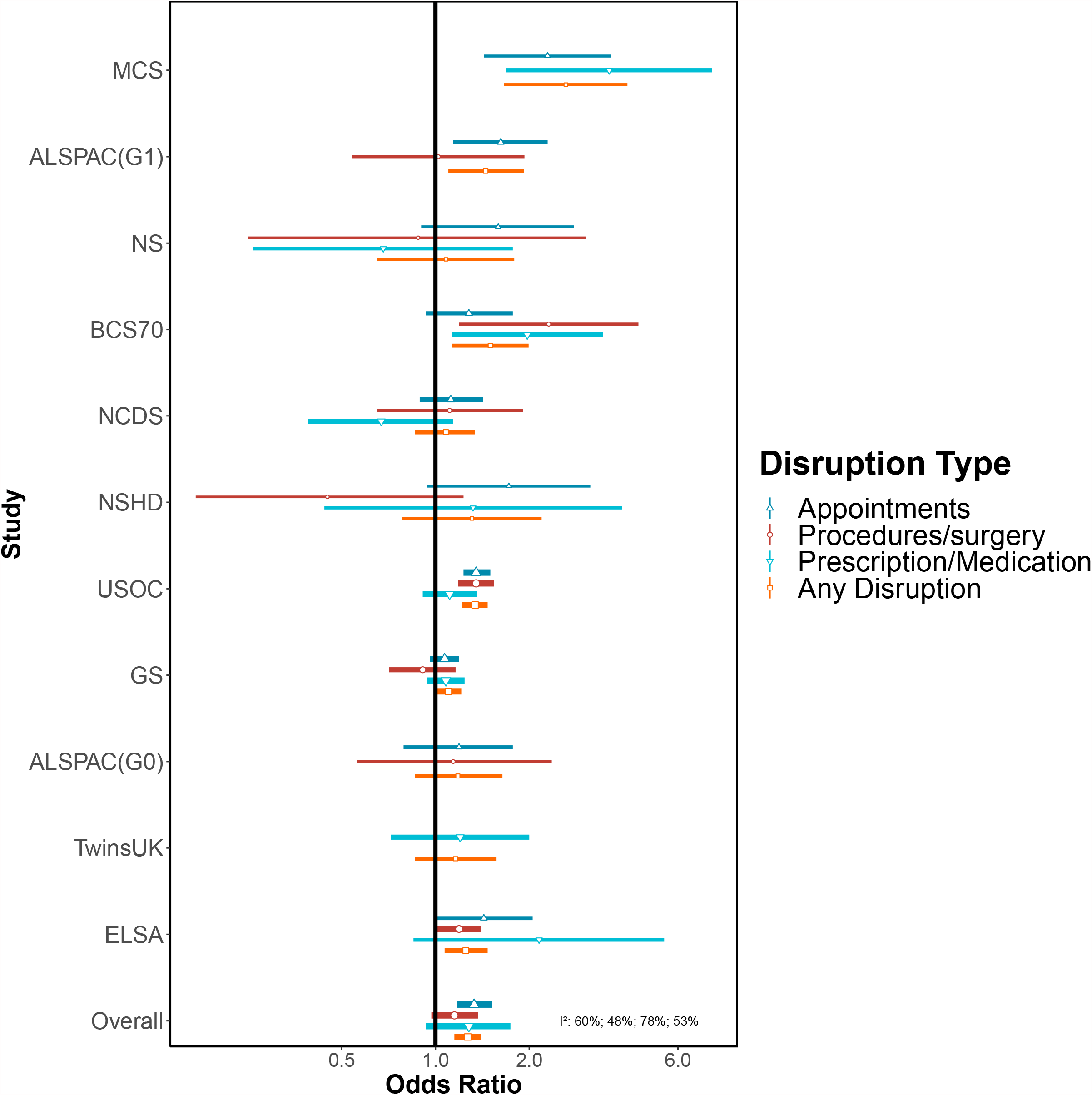
Associations between female (compared to male) sex and healthcare disruption. Notes: Adjusted for age and ethnicity where applicable.

When stratified by age, the odds of having any healthcare disruption for females was highest among 16–24-year-olds (OR: 2.22 [95% CI 1.63, 3.02]; I^2^=0%, Supplementary Table S4). An association between sex and healthcare disruption was observed up to age 54 years but there were no clear associations among those aged 55 years and above. In both the shielding and non-shielding groups, females were more likely to report healthcare disruption (Supplementary Table S5).

### Age and healthcare disruptions

A higher prevalence of having any healthcare disruption was observed among older participants of the national birth cohorts where the same questionnaire was used (Figure 1). This age difference was also observed among the ALSPAC studies and for other age-heterogenous studies as seen in Supplementary Table S3.

The meta-analysis of age-heterogenous studies were supportive of age differences for any healthcare disruptions e.g., OR: 1.39 [1.13,1.72]; I^2^=77% for 65-75y vs 45-54y (Figure 3, Supplementary File 2). Disruptions seemed less likely in younger age groups and more likely among older age groups, though some estimates cross the null and had high heterogeneity, which may be because of few studies in specific age categories (Figure 3, Supplementary File 2). Associations for disruptions to medical appointments and procedures or surgery showed these age differences more clearly (Figure 3, Supplementary File 2).

**Figure 3:**
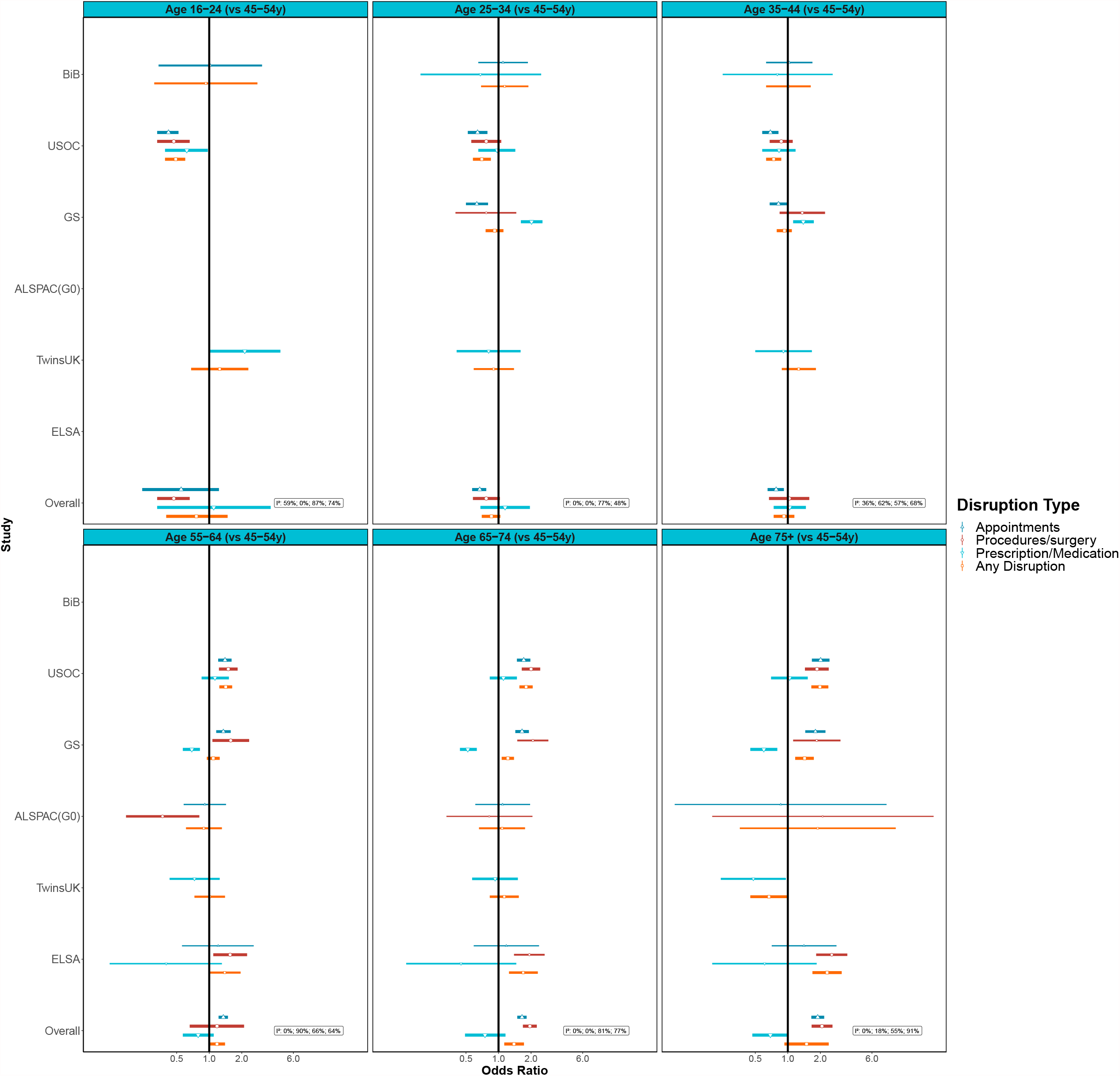
Associations between age (compared to 45-54 year olds) and healthcare disruption. Notes: Adjusted for sex and ethnicity where applicable.

Associations with age and any healthcare disruption were clearest among those not shielding. For those who were shielding, confidence intervals were wide and over-lapped both the null and the estimates from the non-shielding population (Supplementary Table S5). The magnitude for the association of healthcare disruption among 75-year-olds and above vs 45-54 year olds was higher among the non-shielding group (OR: 1.61 [95%CI: 1.17,2.22]; I^2^=79%) compared with the shielding group (OR: 0.83 [95%CI:0.51,1.37; I^2^=33% Supplementary Table S5).

### Ethnicity and healthcare disruptions

Among the studies that had an ethnically diverse sample, between 7.8% (BIB) and 31.9% (USOC) of the White groups reported healthcare disruption. Between 8.3% (TWINSUK) and 23.6% (GS) of Ethnic minority groups reported having any healthcare disruption (Supplementary Table S3).

In meta-analysis, Ethnic minorities compared to White groups had increased odds of any healthcare disruption (OR: 1.19 [1.05,1.35]; I^2^=0%, Figure 4 and Supplementary File 2). This association was less clear for specific domains of healthcare disruption (Figure 4, Supplementary File 2). Among the studies that had a finer categorisation of ethnicity, only the Black ethnic groups had clearly raised odds for any healthcare disruption compared with White groups (OR: 1.38 [1.03,1.84]; I^2^=0%). Associations with healthcare disruption were less evident for other ethnic groups but were imprecisely estimated (Figure 4, Supplementary File 2).

**Figure 4:**
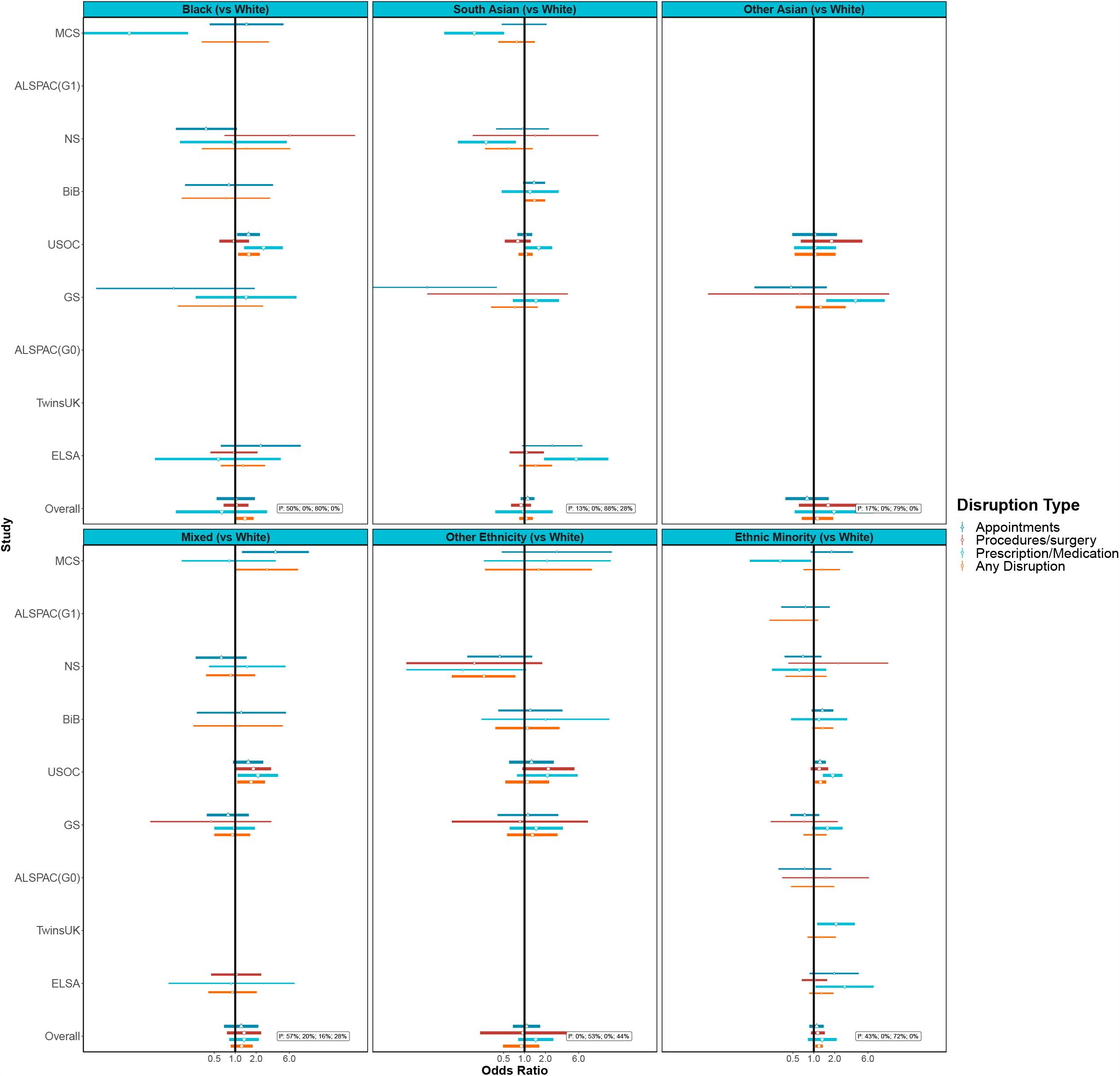
Associations between Ethnicity (compared to White groups) and healthcare disruption. Notes: Panels illustrate findings for some larger ethnic groups separately and the final panel presents results for all non-White ethnic minorities combined. Adjusted for age and sex where applicable.

Associations between ethnicity and any healthcare disruption were not evident within many age bands, though this may simply be due to low power as confidence intervals were wide (Supplementary Table S4). The clearest associations with Ethnic minority groups were within the 35-44- and 45-74-year age ranges (OR:1.31 [1.01,1.71]; I^2^=0% and OR:1.61 [1.16,2.22]; I^2^=0%). The mixed ethnicity group were also at particular risk for disruption in the 16–24-year age range too (OR:2.50 [1.25,5.02]; I^2^=0%). The magnitude for the association between any healthcare disruption among Ethnic minority groups vs. White groups was higher among those who were shielding (OR: 1.56[1.01 to 2.39]; compared to OR: 1.06[0.86 to 1.31] for non-shielding). This observation was consistent across more granular ethnicity categories, but confidence intervals were wide (Supplementary Table S5).

### Education and healthcare disruptions

There was no clear pattern in the prevalence of healthcare disruption across education levels. For example, in USOC 29.7% of those with any healthcare disruption had a degree or equivalent and 39% had no school-leaving qualifications. In TWINSUK 9.9% of those with any healthcare disruption had a degree or equivalent and 6.1% had no school-leaving (Supplementary Table S3).

In meta-analysis, we did not observe clear associations between education level and healthcare disruption, other than that those without school-leaving qualifications had raised odds of disruptions to procedures or surgery (OR: 1.26 [1.11,1.44]; I^2^=0%; Supplementary File 2 and Figure 5). Associations for not having school-leaving qualifications were particularly evident in the 45–74-year age range but had wide confidence intervals at other ages (Supplementary Table S4). We did not observe differences by shielding status (Supplementary Table S5).

**Figure 5:**
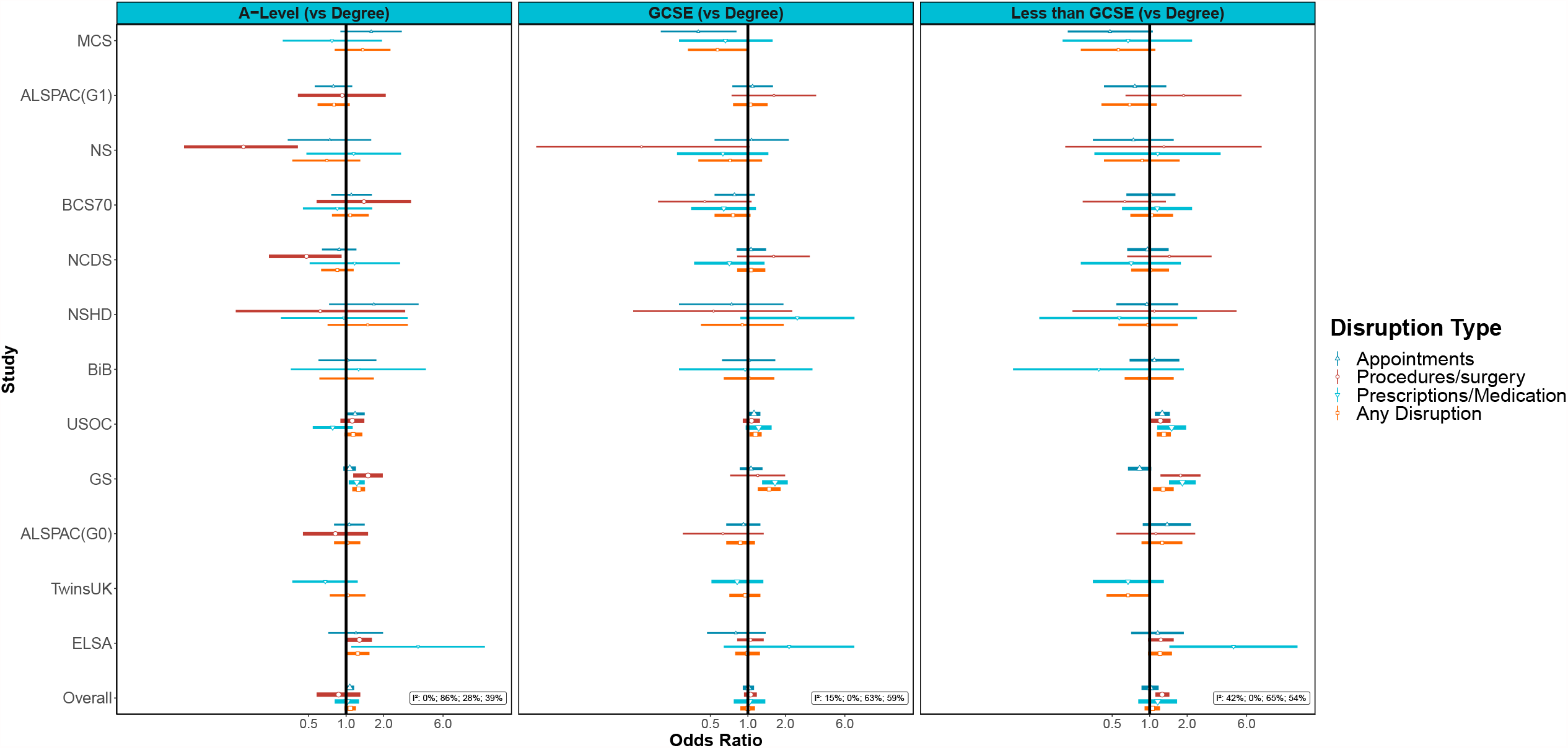
Associations between education (compared to degree level) and healthcare disruption. Notes: Adjusted for age, sex and ethnicity where applicable.

### Occupational class and healthcare disruptions

The prevalence of any healthcare disruption ranged between 9.7% (BIB) and 25.7% (USOC) among the Professional/Managerial social class and between 9.3% (BIB) and 27.6% (USOC) for the Manual/Routine social class (Supplementary Table S3).

Results from meta-analysis show that those in a more disadvantaged occupational class were more likely to report any healthcare disruptions (e.g., OR: 1.17 [95%CI: 1.08, 1.27]; I^2^=0% for Manual/Routine compared with Professional/Managerial, Figure 6, Supplementary File 2). The OR was greatest for the non-employed occupational class category (OR: 1.51 [95%CI: 1.12,2.40]), however the I^2^ was also large (80%). This implies considerable between study heterogeneity, though two of the three individual studies (MCS and ELSA) that did not show clear associations for this category were at the extremes of the age range considered. Similar associations were seen for domains of healthcare disruption, with the largest inequalities seen for access to medications.

**Figure 6:**
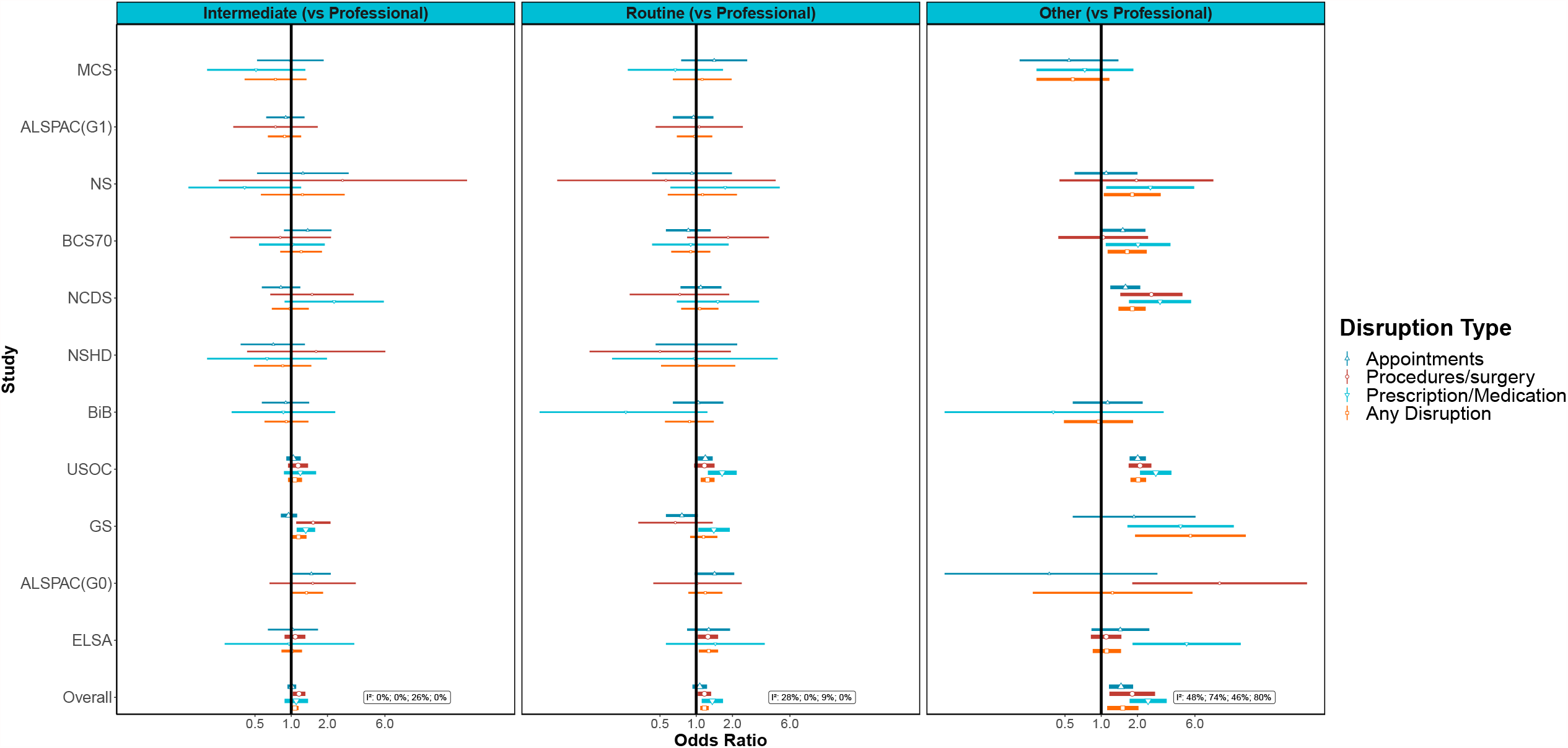
Associations between occupational social class (compared to Professional/Managerial) and healthcare disruption. Notes: Adjusted for age, sex and ethnicity where applicable.

The associations with occupational class for both routine/manual occupations and long-term non-employment were most clearly evident from ages 25-64, i.e., primarily among working age participants (Supplementary Table S4). Associations with occupational class were clearest among those not shielding. Among those who were shielding the associations with occupational class were less evident, but confidence intervals were wide and over-lapped both the null and the estimates from the non-shielding population (Supplementary Table S5).

### Further Adjustment

The above associations for sex, age and long-term non-employment were largely robust to further adjustment for education, occupational class, UK Nation (where appropriate), household composition, and pre-pandemic self-reported health (see Supplementary File 2).

## Discussion

Our study demonstrates marked inequalities in healthcare disruption during the COVID-19 pandemic by harnessing data from 12 UK longitudinal studies. Females were more likely to report healthcare disruptions than males, especially at younger ages (<55 years). This inequality was observed for each healthcare disruption type including prescription medication, procedures or surgery, and appointments as well as a combined measure for any of these disruptions. Older adults were especially likely to report disruptions to medical appointments and procedures and surgeries compared to their younger counterparts. Ethnic minority (excluding White minorities) groups were more likely to report healthcare disruption compared to White (including White minorities) groups. Furthermore, when stratifying results by shielding status, the magnitude for the association between any healthcare disruption among Ethnic minority groups (compared to White groups) was higher among those who were shielding. In studies where a finer breakdown of ethnicity was possible, Black ethnic minority groups had the most clearly increased odds of disruption compared to White ethnic groups. Occupational class was also found to be associated with healthcare disruption with those in a routine/manual occupation or other (which included never/long-term non-employed) being more likely to experience healthcare disruption than those in a managerial/professional occupation, and this inequality appeared to be concentrated within working ages (i.e. 25-64 years). No clear association between education and healthcare disruption was found in the main, age or shielding status stratified analyses.

The direct burden of COVID-19 on health services across the globe has been colossal and remains so in some countries, with prioritisation of COVID-19 patients, leaving less capacity and resources for non-COVID-19 healthcare. Furthermore, associated repeated lockdown measures are also likely to decrease healthcare access and availability with a decrease in the number of people attending A&E services,^4^ and reports of difficulties accessing medication.^8^

Our findings are consistent with current evidence from a smaller sub-set of the studies examined here suggesting that females are more likely to experience disruption to planned surgery, medical procedures, or other medical appointments during lockdown.^12^

Furthermore, our results show that older adults were more likely to report healthcare disruption as compared to their younger counterparts, especially disruptions to medical appointments and planned procedures or surgeries. This finding is consistent with current UK evidence indicating that older adults experience more delays and disruption to health services.^32–35^

Black ethnic minority groups were also found to be at increased risk of healthcare disruption compared to white ethnic groups – an issue of particular concern given pre-pandemic ethnic inequalities in healthcare.^36^ The inequalities by occupational class we found are consistent with prior evidence of socioeconomic healthcare inequalities reported in the UK in the past decade,^37^ and highlight that these have still been present in the COVID-19 pandemic. Associations with occupational class were clearer than those for education, which is also an indicator of socioeconomic position but may have been a more distal influence.

## Strengths and limitations

The analysis brings together data from 12 longitudinal studies with rich and sensitive information on healthcare disruption. This study is strengthened by the coordinated investigation in multiple longitudinal studies with differing study designs, different target populations, and varying selection and attrition processes. Moreover, the use of multiple studies increased statistical power to look at subpopulations such as ethnic minority groups across cohorts and allowed for greater examination of how inequalities were patterned by age. Our novel approach to coordinated analyses harnessing multiple datasets therefore allowed research questions to be addressed which would not otherwise be possible.

Differences between studies in a range of factors including measurement of healthcare disruption, timing of surveys, design, response rates, and differential selection into the COVID-19 sweeps are potentially responsible for heterogeneity in estimates. However, despite this heterogeneity, the key findings were consistent across most datasets. Furthermore, this heterogeneity can be informative, for example, by virtue of mixing age-specific and age range studies, we identified that sex inequalities were stronger at younger ages, and inequalities by occupational class were concentrated within working ages. The definition of healthcare disruption used may also have contained a range of disruptions of greater or lesser severity, and there may have been further inequalities in the severity of disruptions experienced.

We have focused on our aim of identifying who experienced greater disruptions in healthcare, rather than on adjustment for confounders to estimate causal effects of the exposures in question.^38^ Nevertheless, many of the associations we observed were robust to adjustment for a wider range of related variables, but bias due to residual confounding cannot be ruled out. Importantly, we did not condition our analyses on healthcare need. Many of the inequalities we observed for healthcare disruptions may be due to inequalities in health, with those who have greater health needs being more likely to require healthcare that could be disrupted. Accounting for differences in need could have masked inequalities in healthcare disruptions that are caused by inequalities in health and could have made it less clear which groups have been more likely to experience disruption during the pandemic. Restricting analyses to those who needed care could also induce bias if there were unmeasured determinants of both need and disruption.^39^ Nevertheless, another study of the USOC data analysed here that did restrict analyses to those needing care still found income-related inequalities in healthcare disruption, and most of the associations we observed were robust to adjustment for pre-pandemic self-assessed health.^40^

### Impact of healthcare disruption

Disadvantaged groups such as females, older adults, Black ethnic minority groups, and those in routine/manual occupations have had elevated odds of healthcare disruption in the first 8-10 months of the COVID-19 pandemic. Delays and disruptions to treatment could have ongoing implications for patients’ physical and mental health.^41^ Action is needed to remedy these inequalities, and efforts to ensure continuity of care during pandemic-related disruptions may need to be more clearly targeted to those who most need that care. As healthcare access resumes, given the forgone delays in treatments and the subsequent backlog of postponed surgeries,^42^ these groups may require prioritised support to address unmet needs experienced during the pandemic.

### Conclusion

There have been clear inequalities in disruptions to healthcare during the COVID-19 pandemic in the UK. Females (especially at younger ages), older adults, ethnic minorities, and those in disadvantaged occupational classes (especially at working ages) have been more likely to experience healthcare disruptions. These are groups who usually experience worse health, so disruptions related to COVID-19 have clear potential to maintain or even exacerbate existing health inequalities.

## Supporting information

Supplementary file 1 -- Variable coding

Supplementary file 2 -- MATables

## Data Availability

Data for NCDS (SN 6137), BCS70 (SN 8547), Next Steps (SN 5545), MCS (SN 8682) and all four COVID-19 surveys (SN 8658) are available through the UK Data Service. NSHD data are available on request to the NSHD Data Sharing Committee. Interested researchers can apply to access the NSHD data via a standard application procedure. Data requests should be submitted to; further details can be found at http://www.nshd.mrc.ac.uk/data.aspx. doi:10.5522/NSHD/Q101; doi:10.5522/NSHD/Q10. ALSPAC data is available to researchers through an online proposal system. Information regarding access can be found on the ALSPAC website (http://www.bristol.ac.uk/media-library/sites/alspac/documents/researchers/data-access/ALSPAC_Access_Policy.pdf). The TwinsUK Resource Executive Committee (TREC) oversees management, data sharing and collaborations involving the TwinsUK registry (for further details see https://twinsuk.ac.uk/resources-for-researchers/access-our-data/). Understanding Society data are available through the UK Data Service (SN 6614 and SN 8644). Waves 1-9 of ELSA are available through the UK Data Service (SN 8688 and 5050). Access to Generation Scotland data is approved by the Generation Scotland Access Committee. See https://www.ed.ac.uk/generation-scotland/for-researchers/access or for further details. Born in Bradford data is available to researchers through an Expression of Interest form which is reviewed by the Executive group on a monthly basis. See https://borninbradford.nhs.uk/research/how-to-access-data/.

## People acknowledgments

GS: Drew Altschul, Chloe Fawns-Ritchie, Archie Campbell, Robin Flaig.

ALSPAC: Daniel J Smith, Nicholas J Timpson, Kate Northstone.

Understanding Society: Michaela Benzeval

TwinsUK: Deborah Hart, María Paz García, Rachel Horsfall, Ruth C.E. Bowyer.

CLS: Matt Brown, Lisa Calderwood, Emla Fitzsimons, Alissa Goodman, Aida Sanchez

NSHD: Andrew Wong, Maria Popham, Karen MacKinnon, Imran Shah, Philip Curran

## Notes

### Competing Interest Statement

Srinivasa Vittal Katikireddi is a member of the Scientific Advisory Group on Emergencies subgroup on ethnicity and COVID-19 and is co-chair of the Scottish Government Ethnicity Reference Group on COVID-19. Nish Chaturvedi serves on a data safety monitoring board for trials sponsored by Astra-Zeneca.

### Funding Statement

Funding acknowledgements
This work was supported by the National Core Studies, an initiative funded by UKRI, NIHR and the Health and Safety Executive. The COVID-19 Longitudinal Health and Wellbeing National Core Study was funded by the Medical Research Council (MC_PC_20030).
The contributing studies have been made possible because of the tireless dedication, commitment and enthusiasm of the many people who have taken part. We would like to thank the participants and the numerous team members involved in the studies including interviewers, technicians, researchers, administrators, managers, health professionals and volunteers. We are additionally grateful to our funders for their financial input and support in making this research happen.
Studies funding:
Understanding Society is an initiative funded by the Economic and Social Research Council and various Government Departments, with scientific leadership by the Institute for Social and Economic Research, University of Essex, and survey delivery by NatCen Social Research and Kantar Public. The Understanding Society COVID-19 study is funded by the Economic and Social Research Council (ES/K005146/1) and the Health Foundation (2076161). The research data are distributed by the UK Data Service.
The Millennium Cohort Study, Next Steps, British Cohort Study 1970 and National Child Development Study 1958 are supported by the Centre for Longitudinal Studies, Resource Centre 2015-20 grant (ES/M001660/1) and a host of other co-funders. The 1946 NSHD cohort is hosted by the MRC Unit for Lifelong Health and Ageing funded by the Medical Research Council (MC_UU_00019/1Theme 1: Cohorts and Data Collection). The COVID-19 data collections in these five cohorts were funded by the UKRI grant Understanding the economic, social and health impacts of COVID-19 using lifetime data: evidence from 5 nationally representative UK cohorts (ES/V012789/1).
The English Longitudinal Study of Ageing was developed by a team of researchers based at University College London, NatCen Social Research, the Institute for Fiscal Studies, the University of Manchester and the University of East Anglia. The data were collected by NatCen Social Research. The funding is currently provided by the National Institute on Aging in the US, and a consortium of UK government departments coordinated by the National Institute for Health Research. Funding has also been received by the Economic and Social Research Council. The English Longitudinal Study of Ageing Covid-19 Substudy was supported by the UK Economic and Social Research Grant (ESRC) ES/V003941/1.
The UK Medical Research Council and Wellcome (Grant Ref: 217065/Z/19/Z) and the University of Bristol provide core support for ALSPAC. A comprehensive list of grants funding is available on the ALSPAC website (http://www.bristol.ac.uk/alspac/external/documents/grant-acknowledgements.pdf). We are extremely grateful to all the families who took part in this study, the midwives for their help in recruiting them, and the whole ALSPAC team, which includes interviewers, computer and laboratory technicians, clerical workers, research scientists, volunteers, managers, receptionists, and nurses. Please note that the study website contains details of all the data that is available through a fully searchable data dictionary and variable search tool" and reference the following webpage: http://www.bristol.ac.uk/alspac/researchers/our-data/. Ethical approval for the study was obtained from the ALSPAC Ethics and Law Committee and the Local Research Ethics Committees.
TwinsUK receives funding from the Wellcome Trust (WT212904/Z/18/Z), the National Institute for Health Research (NIHR) Biomedical Research Centre based at Guy's and St Thomas' NHS Foundation Trust and King's College London. TwinsUK is also supported by the Chronic Disease Research Foundation and Zoe Global Ltd. The funders had no role in study design, data collection and analysis, decision to publish, or preparation of the manuscript.
Generation Scotland received core support from the Chief Scientist Office of the Scottish Government Health Directorates [CZD/16/6] and the Scottish Funding Council [HR03006]. Genotyping of the GS:SFHS samples was carried out by the Genetics Core Laboratory at the Wellcome Trust Clinical Research Facility, Edinburgh, Scotland and was funded by the Medical Research Council UK and the Wellcome Trust (Wellcome Trust Strategic Award: STratifying Resilience and Depression Longitudinally (STRADL) Reference 104036/Z/14/Z). Generation Scotland is funded by the Wellcome Trust (216767/Z/19/Z).
Born in Bradford (BiB) receives core infrastructure funding from the Wellcome Trust (WT101597MA), and a joint grant from the UK Medical Research Council (MRC) and UK Economic and Social Science Research Council (ESRC) (MR/N024397/1) and one from the British Heart Foundation (BHF) (CS/16/4/32482). The National Institute for Health Research Yorkshire and Humber ARC, and Clinical Research Network both provide support for BiB research.
People funding:
SVK acknowledges funding from a NRS Senior Clinical Fellowship (SCAF/15/02), the Medical Research Council (MC_UU_00022/2) and the Scottish Government Chief Scientist Office (SPHSU17). ASFK acknowledges funding from the ESRC (ES/V011650/1). DJP acknowledges funding from the Wellcome Trust (216767/Z/19/Z and 221574/Z/20/Z). EJT acknowledges funding from the Wellcome Trust (WT212904/Z/18/Z). GBP acknowledges funding from the Economic and Social Research Council (ES/V012789/1).

### Author Declarations

Ethical approval was obtained from the ALSPAC Ethics and Law Committee and the Local Research Ethics Committees. Ethical approval for Born in Bradford was granted by the National Health Service Health Research Authority Yorkshire and the Humber (Bradford Leeds) Research Ethics Committee (reference: 16/YH/0320). The University of Essex Ethics Committee has approved all data collection for the Understanding Society main study and COVID-19 waves. No additional ethical approval was necessary for this secondary data analysis. Waves 1-9 of ELSA were approved through the National Research Ethics Service, while the COVID-19 Sub-study was approved by the UCL Research Ethics Committee. All participants provided informed consent. Generation Scotland obtained ethical approval from the East of Scotland Committee on Medical Research Ethics (on behalf of the National Health Service). Reference number 20/ES/0021. All wave of TwinsUK have received ethical approval associated with TwinsUK Biobank (19/NW/0187), TwinsUK (EC04/015) or Healthy Ageing Twin Study (H.A.T.S) (07/H0802/84) studies from NHS Research Ethics Committees at the Department of Twin Research and Genetic Epidemiology, KCL.

