## Supplementary Tables for "Inequalities in healthcare disruptions during the Covid-19 pandemic: Evidence from 12 UK population-based longitudinal studies"

**List of Supplementary Tables**

**Supplementary Table S1***.* Details of each study

#### **Supplementary Table S2.** Ethics and data access statements for each study

#### **Supplementary Table S3**. Percent prevalence of any healthcare disruptions by selected characteristics and study

#### **Supplementary Table S4.** Meta-analysed associations between each predictor and overall healthcare disruption stratified by age

### **Supplementary Table S5.** Meta-analysed associations between each predictor and overall healthcare disruption stratified by shielding status

**Supplementary Table S1**. Details of each study

| **Study Population** | **Design and Sample Frame** | **2020 Age Range** | **Pre-pandemic Survey** | **Details of Covid surveys**  **(response rate)** | **Analytic N** |
| --- | --- | --- | --- | --- | --- |
| *Age Homogenous Cohorts* | |  |  |  |  |
| MCS: Millennium Cohort Study | Cohort of UK children born between Sept 2000 and Jan 2002 with regular follow-up surveys from birth. | 18-20 | 2018 | Two surveys: May (26.6%) & Sep-Oct (24.2%) | 3147 |
| ALSPAC (G1): Avon Longitudinal Study of Parents and Children- Generation 1 | Cohort of children born in the South-West of England between April 1991 and Dec 1992, with regular follow-up surveys from birth.  (original young people) | 27-29 | 2017-2018 | Three questionnaires: April (19%), June (17.4%), December (26.4%) | 3430 |
| NS: Next Steps, formerly known as Longitudinal Study of Young People in England | Sample recruited via secondary schools in England at around age 13 with regular follow-up surveys thereafter. | 29-31 | 2015 | Two surveys: May (20.3%) & Sep-Oct (31.8%) | 3311 |
| BCS70: British Cohort Study 1970 | Cohort of all children born in Great Britain (i.e. England, Wales & Scotland) in one week in 1970, with regular follow-up surveys from birth. | 50 | 2016 | Two surveys: May (40.4%) & Sep-Oct (43.9%) | 5175 |
| NCDS: National Child Development Study | Cohort of all children born in Great Britain (i.e. England, Wales & Scotland) in one week in 1958, with regular follow-up surveys from birth. | 62 | 2013 | Two surveys: May (57.9%) & Sep-Oct (53.9%) | 5747 |
| NSHD: National Survey of Health and Development | Cohort of all children born in Great Britain (i.e. England, Wales & Scotland) in one week in 1946, with regular follow-up surveys from birth. | 74 | 2015 | Two surveys: May (68.2%) & Sep-Oct (61.5%) | 1569 |
| *Age Heterogeneous Studies* | |  |  |  |  |
| BIB: Born in Bradford | Birth cohort recruiting pregnant women and their children between 2007 and 2010; and pregnant women and their children in three deprived areas of Bradford between 2016 and 2020 | 17-54 | 2016-2020 | Two surveys: April-Jun (28%) & Oct-Nov (24%) | 1726 |
| USOC: Understanding Society: the UK Household Longitudinal Survey | A nationally representative longitudinal household panel study, based on a clustered-stratified probability sample of UK households, with all adults aged 16+ in chosen households surveyed annually. | 16-96 | 2018-2019 | Six: surveys: April (40.3%); May (33.6%); Jun (32.0%); July (31.2%); Sep (29.2%) & Nov (27.3%) | 13253 |
| ELSA: English Longitudinal Study of Aging | A nationally-representative population study of individuals aged 50+ living in England, with biennial surveys since 2002/03. | 52-90+ | 2018-2019 | First Covid-19 sub-study: Jun-July (75%) | 6508 |
| GS: Generation Scotland: the Scottish Family Health Study | A family-structured, population-based Scottish cohort, with participants aged 18-99 recruited between 2006-2011 | 27-100 | 2006-2011 | Two surveys: April-Jun (21.6%) & Jul-Aug (15.6%) | 17139 |
| ALSPAC(G0): Avon Longitudinal Study of Parents and Children- Generation 0 | Parents of the ALSPAC(G1) cohort described above, treated as a separate age-heterogenous study population.  (original parents) | 45-81 | 2011-2013 | Three questionnaires: April (12.4%), June (12.2%), December (14.3%) | 3625 |
| TWINSUK: the UK Adult Twin Registry | A cohort of UK volunteer adult twins (55% monozygotic and 43% dizygotic) who were sampled between 18-101 years of age. | 22-96 | 2017-2018 | Three surveys: April (64.3%), July (77.6%) & November (76.1%) | 4282 |

#### **Supplementary Table S2.** Ethics and data access statements for each study

| **NSHD, NCDS, BCS70**, **NS** and **MCS** | The most recent sweeps of the **NSHD, NCDS, BCS70**, **Next Steps** and **MCS** have all been granted ethical approval by the National Health Service (NHS) Research Ethics Committee and all participants have given informed consent. Data for NCDS (SN 6137), BCS70 (SN 8547), Next Steps (SN 5545), MCS (SN 8682) and all four COVID-19 surveys (SN 8658) are available through the UK Data Service. NSHD data are available on request to the NSHD Data Sharing Committee. Interested researchers can apply to access the NSHD data via a standard application procedure. Data requests should be submitted to; further details can be found at <http://www.nshd.mrc.ac.uk/data.aspx>. doi:10.5522/NSHD/Q101; doi:10.5522/NSHD/Q10. |
| --- | --- |
| **ALSPAC** | Ethical approval was obtained from the **ALSPAC** Ethics and Law Committee and the Local Research Ethics Committees. The study website contains details of all the data that is available through a fully searchable data dictionary and variable search tool: <http://www.bristol.ac.uk/alspac/researchers/our-data>. ALSPAC data is available to researchers through an online proposal system. Information regarding access can be found on the ALSPAC website (<http://www.bristol.ac.uk/media-library/sites/alspac/documents/researchers/data-access/ALSPAC_Access_Policy.pdf>). |
| **BIB** | Ethical approval for **Born in Bradford** was granted by the National Health Service Health Research Authority Yorkshire and the Humber (Bradford Leeds) Research Ethics Committee (reference: 16/YH/0320). Data from the various BiB family studies are available to researchers; see the study website for information on how to access data (<https://borninbradford.nhs.uk/research/how-to-access-data/>). |
| **USOC** | The University of Essex Ethics Committee has approved all data collection for the **Understanding Society** main study and COVID-19 waves. No additional ethical approval was necessary for this secondary data analysis. All data are available through the UK Data Service (SN 6614 and SN 8644). |
| **ELSA** | Waves 1-9 of **ELSA** were approved through the National Research Ethics Service, while the COVID-19 Sub-study was approved by the UCL Research Ethics Committee. All participants provided informed consent. All data are available through the UK Data Service (SN 8688 and 5050). |
| **GS** | **Generation Scotland** obtained ethical approval from the East of Scotland Committee on Medical Research Ethics (on behalf of the National Health Service). Reference number 20/ES/0021. Access to data is approved by the Generation Scotland Access Committee. See <https://www.ed.ac.uk/generation-scotland/for-researchers/access> or for further details. |
| **TWINSUK** | All wave of **TwinsUK** have received ethical approval associated with TwinsUK Biobank (19/NW/0187), TwinsUK (EC04/015) or Healthy Ageing Twin Study (H.A.T.S) (07/H0802/84) studies from NHS Research Ethics Committees at the Department of Twin Research and Genetic Epidemiology, King’s College London. The TwinsUK Resource Executive Committee (TREC) oversees management, data sharing and collaborations involving the TwinsUK registry (for further details see <https://twinsuk.ac.uk/resources-for-researchers/access-our-data/>). |

### **Supplementary Table S3.** Percent prevalence of any healthcare disruptions by selected characteristics and study

|  |  | ***MCS*** | ***ALSPAC***  ***(G1)*** | ***NS*** | | ***BCS70*** | | ***NCDS*** | | ***NSHD*** | ***BIB*** | ***USOC*** | | ***GS*** | | ***ALSPAC***  ***(G0)*** | | ***TWINS UK*** | | ***ELSA*** | |
| --- | --- | --- | --- | --- | --- | --- | --- | --- | --- | --- | --- | --- | --- | --- | --- | --- | --- | --- | --- | --- | --- |
| **Sex** | *Male* | 6.1 | 12.8 | 12.1 | | 11.7 | | 15.6 | | 14.5 | NA | 29.4 | | 24.9 | | 18.1 | | 7.4 | | 17.5 | |
|  | *Female* | 14.1 | 17.5 | 13.8 | | 16.9 | | 17.4 | | 18.2 | 9.4 | 34.0 | | 25.5 | | 20.5 | | 8.5 | | 21.3 | |
| **Age** | *16-24* | 10.1 |  |  | |  | |  | |  | 8.2 | 18.3 | | NA | | NA | | 10 | | NA | |
|  | *25-34* |  | 15.9 | 12.8 | |  | |  | |  | 10.4 | 24.0 | | 22.9 | | NA | | 7.7 | | NA | |
|  | *35-44* |  |  |  | |  | |  | |  | 9.1 | 24.9 | | 23.0 | | NA | | 13.2 | | NA | |
|  | *45-54* |  |  |  | | 14.3 | |  | |  | 8.7 | 30.9 | | 24.2 | | 21.3 | | 13.9 | | 13.0 | |
|  | *55-64* |  |  |  | |  | | 16.7 | |  | -- | 38.6 | | 25.2 | | 19.2 | | 21.6 | | 17.2 | |
|  | *65-74* |  |  |  | |  | |  | | 16.4 | -- | 43.6 | | 26.8 | | 21.8 | | 31.4 | | 20.0 | |
|  | *75+* |  |  |  | |  | |  | |  | -- | 45.6 | | 29.2 | | 30.6 | | 9.2 | | 25.5 | |
| **Ethnicity** | *White* | 10.0 | 16.1 | 13.3 | | -- | | -- | | -- | 7.8 | 31.9 | | 25.4 | | 19.9 | | 8.3 | | 19.5 | |
|  | *South Asian* | 6.6 | NA | 8.4 | | -- | | -- | | -- | 10.6 | 25.4 | | 20.0 | | NA | | 5.1 | | 22.9 | |
|  | *OtherAsian* | NA | NA | NA | | -- | | -- | | -- | NA | 37.5 | | 27.4 | | NA | | 11.1 | | NA | |
|  | *Black* | 7.7 | NA | 18.8 | | -- | | -- | | -- | 5.9 | 35.8 | | 19.0 | | NA | | 11.5 | | 21.7 | |
|  | *Mixed* | 23.5 | NA | 11.1 | | -- | | -- | | -- | 8.3 | 27.7 | | 22.9 | | NA | | 10 | | 15.5 | |
|  | *Other* | 11.1 | NA | 4.2 | | -- | | -- | | -- | 8.5 | 30.2 | | 28.6 | | NA | | 9.1 | | NA | |
|  | *All ethnic Minorities* | 10.6 | 9.0 | 10.7 | | -- | | -- | | -- | 10.3 | 30.4 | | 23.6 | | 19.6 | | 8.3 | | 21.1 | |
| **Education** | *Higher Ed* | 11.2 | 16.9 | 14.0 | | 14.5 | | 16.8 | | 16.03 | 9.0 | 29.7 | | 23.3 | | 19.4 | | 9.9 | | 16.9 | |
|  | *A-level* | 14.8 | 14.4 | 10.5 | | 15.5 | | 14.0 | | 22.67 | 9.2 | 27.0 | | 26.7 | | 20.0 | | 10.3 | | 20.5 | |
|  | *GCSE* | 6.3 | 18.1 | 11.3 | | 12.0 | | 17.6 | | 15.6 | 9.0 | 31.3 | | 29.3 | | 17.8 | | 9.2 | | 17.4 | |
|  | *<GCSE/ None* | 6.2 | 12.4 | 14.5 | | 15.5 | | 17.2 | | 16.3 | 9.1 | 39.0 | | 27.8 | | 23.9 | | 6.1 | | 22.4 | |
| **Social Class** | *Managerial/ Admin/ Professional* | 11.6 | 16.4 | 11.1 | | 12.6 | | 12.7 | | 17.0 | 9.7 | 25.7 | | 24.3 | | 16.4 | | - | | 18.3 | |
|  | *Intermediate* | 8.5 | 15.2 | 12.7 | | 15.3 | | 12.6 | | 15.5 | 9.0 | 27.2 | | 25.7 | | 21.3 | | - | | 19.5 | |
|  | *Manual/Routine* | 11.2 | 16.7 | 11.6 | | 11.6 | | 13.6 | | 18.6 | 9.3 | 27.6 | | 25.6 | | 19.6 | | - | | 23.4 | |
|  | *Other* | 6.0 | 0 | 18.0 | | 19.3 | | 21.1 | | 0 | 11.8 | 42.6 | | 51.9 | | 20.0 | | - | | 16.6 | |
| **Not Instructed to Shield** | | 9.0 | -- | 12.0 | 12.4 | | 14.6 | | 16.7 | | -- | | 29.6 | | 23.9 | | -- | | 8.9 | | 16.2 |
| **Instructed to Shield** | | 47.5 | -- | 44.3 | 49.4 | | 41.9 | | 28.4 | | -- | | 61.0 | | 42.0 | | -- | | 15.3 | | 35.5 |

Sources: MCS (Millennium Cohort Study); ALSPAC G1 (Children of the Avon Longitudinal Study of Parents and Children); NS (Next Steps); BCS 70 (1970 British Cohort Study), NCDS (National Child Development Study); NSHD (National Survey of Health and Development); BIB (Born in Bradford); ; USOC (Understanding Society); GS (Generation Scotland: the Scottish Family Health Study); ALSPAC G0 (parents of ALSPAC); TWINS UK (UK Adult Twin Registry); ELSA (English Longitudinal Study of Ageing). Notes: Samples for each study restricted to respondents with non-missing information on healthcare disruptions and valid information on sex, social class, education and (where applicable) age and ethnicity. All information about how information was collected and variables were coded is available in Supplementary File 1. NA= Not available; (--)= Info not collected. Weighted data where applicable

### **Supplementary Table S4.** Meta-analysed associations between each predictor and overall healthcare disruption stratified by age

|  |  | **Any Healthcare disruption** | | | | | | |
| --- | --- | --- | --- | --- | --- | --- | --- | --- |
|  |  | **16-24** | **25-34** | **35-44** | **45-54** | **55-64** | **65-74** | **75+** |
|  |  | OR (95% CI) | OR (95% CI) | OR (95% CI) | OR (95% CI) | OR (95% CI) | OR (95% CI) | OR (95% CI) |
| **Sex**  **(ref male)** | *Female* | 2.22 (1.63,3.02) | 1.56 (1.3,1.87) | 1.51 (1.23,1.86) | 1.72 (1.35,2.18) | 1.09 (0.92,1.3) | 0.98 (0.79,1.21) | 1.08 (0.90,1.30) |
|  | *% I ^2^* | 0 | 0 | 0 | 36.61 | 59.58 | 70.98 | 20.0 |
| **Ethnicity**  **(ref White)** | *Non-white* | 1.3 (0.89,1.89) | 0.92 (0.65,1.29) | 1.31 (1.01,1.71) | 1.61 (1.16,2.22) | 1.13 (0.85,1.50) | 1.59 (1.14,2.23) | 1.28 (0.67,2.45) |
|  | *% I ^2^* | 0 | 36.48 | 0 | 0 | 0 | 0 | 0 |
|  | *Black* | 1.15 (0.51,2.59) | 0.82 (0.4,1.68) | 1.91 (0.81,4.48) | 1.99 (0.93,4.25) | 1.69 (1,2.84) | 1.16 (0.48,2.8) | 1.23 (0.42,3.56) |
|  | *% I ^2^* | 0 | 0 | 0 | 15.25 | 0 | 0 | 0 |
|  | *Other Asian* | - | 0.62 (0.2,1.92) | 1.63 (0.8,3.32) | 1.75 (0.54,5.64) | 0.96 (0.43,2.15) | 1.10 (0.27, 4.55) | - |
|  | *% I ^2^* | - | 0 | 0 | 0 | 0 | 0 | - |
|  | *South Asian* | 0.98 (0.62,1.53) | 0.8 (0.38,1.71) | 1.11 (0.8,1.55) | 1.67 (0.43,6.48) | 0.82 (0.44,1.56) | 1.55 (0.83,2.9) | 1.11 (0.4,3.12) |
|  | *% I ^2^* | 13.95 | 74.73 | 10.10 | 82.00 | 14.81 | 24.24 | 0 |
|  | *Mixed* | 2.50 (1.25,5.02) | 1.26 (0.79,2.02) | 1.15 (0.23,5.69) | 0.92 (0.46,1.87) | 1.06 (0.53,2.11) | 1.63 (0.7,3.80) | 1.47 (0.34,6.42) |
|  | *% I ^2^* | 0 | 0 | 73.12 | 0 | 0 | 0 | 22.46 |
|  | *Other Ethnic Minority* | 0.18 (0,15.35) | 0.80 (0.31,2.08) | 1.41 (0.58,3.40) | 1.74 (0.56,5.45) | 0.77 (0.27,2.22) | - | - |
|  | *% I ^2^* | 88.56 | 49.28 | 0 | 29.75 | 0 | - | - |
| **Education**  **(ref Degree)** | *A-level* | 1.33 (0.93,1.90) | 0.99 (0.69,1.42) | 1.62 (1.28,2.05) | 1.13 (0.96,1.34) | 1.01 (0.89,1.14) | 1.25 (1.10,1.43) | 0.96 (0.65,1.4) |
|  | *% I ^2^* | 0 | 62.16 | 0 | 0 | 0 | 0 | 57.49 |
|  | *GCSE* | 0.94 (0.49,1.81) | 1.24 (0.8,1.94) | 1.26 (0.97,1.63) | 1.16 (0.83,1.62) | 1.03 (0.91,1.17) | 1.05 (0.90,1.23) | 0.92 (0.65,1.30) |
|  | *% I ^2^* | 64.06 | 69.10 | 0 | 62.52 | 0 | 0 | 35.17 |
|  | *No degree* | 0.77 (0.47,1.28) | 0.99 (0.67,1.45) | 1.03 (0.74,1.43) | 1.48 (1.08,2.04) | 1.20 (1.03, 1.41) | 1.23 (1.07,1.42) | 0.96 (0.78,1.20) |
|  | *% I ^2^* | 11.51 | 42.2 | 0 | 34.96 | 0 | 0 | 0 |
| **Social Class**  **(ref Manager/ Profess)** | *Intermediate* | 0.92 (0.60,1.41) | 1.04 (0.84,1.29) | 1.28 (0.92,1.78) | 1.12 (0.94,1.33) | 1.01 (0.86,1.19) | 0.98 (0.83,1.16) | 1.00 (0.76,1.33) |
|  | *% I ^2^* | 0 | 10.79 | 46.81 | 0 | 22.02 | 0 | 0 |
|  | *Routine/manual* | 1.15 (0.77,1.71) | 1.11 (0.8,1.55) | 1.24 (0.95,1.63) | 1.08 (0.9,1.3) | 1.16 (1,1.35) | 1.06 (0.86,1.31) | 1.27 (0.96,1.67) |
|  | *% I ^2^* | 0 | 50.55 | 23 | 0 | 0 | 7.23 | 0 |
|  | *Other* | 1.02 (0.46,2.26) | 1.85 (1.29,2.64) | 1.44 (0.55,3.80) | 2.45 (1.18,5.06) | 1.65 (1.21,2.27) | 1.22 (0.95,1.56) | 1.02 (0.62,1.69) |
|  | *% I ^2^* | 58.35 | 0 | 68.27 | 76.07 | 60.63 | 0 | 0 |

*Note.* Models adjusted for sex and ethnicity

### **Supplementary Table S5.** Meta-analysed associations between each predictor and overall healthcare disruption stratified by shielding status

|  |  | **Any Healthcare disruption** | |
| --- | --- | --- | --- |
|  |  | **Shielding (no)** | **Shielding (yes)** |
|  |  | OR (95% CI) | OR (95% CI) |
| **Sex**  **(ref male)** | Female | 1.26 (1.12,1.43) | 1.32 (1.1,1.58) |
|  | *% I ^2^* | 61.12 | 0 |
| **Age**  **(ref 45-54)** | 16-24 years | 0.79 (0.4,1.56) | 0.64 (0.23,1.78) |
|  | *% I ^2^* | 70.32 | - * |
|  | 25-34 years | 0.86 (0.7,1.06) | 1.09 (0.61,1.95) |
|  | *% I ^2^* | 43.4 | 0 |
|  | 35-44 years | 0.95 (0.74,1.21) | 0.68 (0.34,1.34) |
|  | *% I ^2^* | 68.26 | 47.41 |
|  | 55-64 years | 1.21 (1.02,1.43) | 1.24 (0.87,1.77) |
|  | *% I ^2^* | 53.82 | 0 |
|  | 65-74 years | 1.44 (1.2,1.72) | 1.11 (0.79,1.56) |
|  | *% I ^2^* | 64.1 | 0 |
|  | 75+ years | 1.61 (1.17,2.22) | 0.83 (0.51,1.37) |
|  | *% I ^2^* | 79.38 | 32.84 |
| **Ethnicity**  **(ref White)** | Non-white | 1.06 (0.86,1.31) | 1.56 (1.01,2.39) |
|  | *% I ^2^* | 41.46 | 0 |
|  | Black | 0.80 (0.43,1.49) | 1.60 (0.67,3.83) |
|  | *% I ^2^* | 58.06 | 0 |
|  | Other Asian | 0.95 (0.54,1.68) | - |
|  | *% I ^2^* | 0 | - |
|  | South Asian | 0.98 (0.75,1.28) | 1.4 (0.83,2.35) |
|  | *% I ^2^* | 35.03 | 0 |
|  | Mixed | 1.18 (0.85,1.62) | 1.85 (0.71,4.77) |
|  | *% I ^2^* | 0 | 0 |
|  | Other Ethnic Minority | 0.85 (0.45,1.62) | 1.47 (0.18,12.14) |
|  | *% I ^2^* | 43.11 | - ** |
| **Education**  **(ref Degree)** | A-level | 1.09 (0.96,1.23) | 0.94 (0.73,1.22) |
|  | *% I ^2^* | 39.28 | 0 |
|  | GCSE | 0.99 (0.84,1.17) | 0.81 (0.62,1.05) |
|  | *% I ^2^* | 64.95 | 0 |
|  | No degree | 1.02 (0.88,1.19) | 0.90 (0.70,1.15) |
|  | *% I ^2^* | 46.14 | 0 |
| **Social Class (ref Managerial / Professional)** | Intermediate | 1.07 (0.98,1.16) | 0.93 (0.71,1.21) |
|  | *% I ^2^* | 0 | 0.60 |
|  | Routine/manual | 1.18 (1.07,1.29) | 0.92 (0.7,1.21) |
|  | *% I ^2^* | 0 | 0 |
|  | Other | 1.48 (1.04,2.09) | 1.48 (0.88,2.52) |
|  | *% I ^2^* | 83.37 | 48.47 |

*Note.* Models adjusted for age and ethnicity ** USOC only **GS only*
